# Structural barriers to social protection and HIV prevention services for sex workers in Southeast Asia: a fixed-effects panel data analysis, 2018–2025

**DOI:** 10.64898/2026.04.12.26350700

**Authors:** Jason Hung, Adrian Smith

**Affiliations:** Department of Sociology and Anthropology, Chulalongkorn University, Bangkok, Thailand; Nuffield Department of Population Health, University of Oxford, Oxford, UK

## Abstract

**Introduction:** Empirical evidence linking the existence of national structural policies to the provision of key HIV services in low- and middle-income settings remains scarce. This study addresses the research gap by quantifying the within-country relationships between six national structural policy indicators and the presence of the HIV prevention service component targeted at sex workers.

**Methods:** We constructed a balanced panel covering eight Southeast Asian countries over 2018 to 2025 from the UNAIDS Global AIDS Monitoring (GAM) framework. For each country and year, the panel records the government’s reported status on six policy indicators, namely (1) any HIV prevention service component targeted at sex workers, (2) barriers to accessing social protection among people living with HIV and key populations, (3) coordination between social protection and the national AIDS programme, (4) national implementation guidelines for HIV prevention programmes for sex workers, (5) a social protection policy addressing unpaid care work, and (6) a social protection policy recognising adolescent girls and young women as key beneficiaries. We used Fixed-Effects (FE) and Random-Effects (RE) models, with the FE model selected as the more statistically appropriate estimator, and one-period lagged explanatory variables.

**Results:** The primary finding from the FE model indicated a statistically significant and positive contemporaneous association between the existence of legal or administrative barriers to social protection and the presence of HIV prevention services for sex workers (*β* = 0.8531; *p* < 0.001). However, the robustness check revealed a statistically significant negative association when using the lagged barrier variable (*β* = −0.3540; *p* = 0.011). No other indicator’s coefficient was statistically significant in the FE models.

**Conclusions:** The sustained presence of these impediments over time is associated with the absence of HIV prevention service provision in the following year. National HIV programmes must urgently prioritise the removal of structural barriers for key populations.

**Key Messages:**

1. **What is already known on this topic:** The global HIV response requires addressing structural determinants, such as legal barriers to social protection, to achieve epidemic control, but there is a lack of robust empirical evidence linking the adoption of specific national structural policies to the actual availability of essential HIV services for key populations in low- and middle-income settings.
2. **What this study adds**: This study provides the first evidence using FE panel data that the existence of national policy barriers to social protection is initially associated with a higher likelihood of reporting an HIV prevention service component for sex workers, and that this positive association is short-lived, with the sustained presence of the barrier associated with the absence of that component in the subsequent year.
3. **How this study might affect research, practice or policy**: Policymakers should recognise that simply identifying and reporting structural barriers is insufficient for sustained policy intervention effectiveness, and policy should focus not just on the adoption of targeted programmes but on the urgent removal of structural barriers for key populations.

## Introduction

The principle of universal access to human immunodeficiency virus (HIV) treatment and prevention has been established in international human rights law over several decades (UNAIDS, 2012). HIV-related needs are also recognised to be greater among key populations, who exert more influence on transmission as a whole. Structural attention to enabling access to treatment and prevention is, therefore, high on the HIV agenda itself. The global HIV response has shifted towards achieving epidemic control through addressing structural determinants that create vulnerability (UNAIDS, 2021), such as access to social protection (Ooms & Kruja, 2019) and mitigating legal barriers to services (Nkengasong & Ratevosian, 2023). Key populations, including sex workers (CATIE, 2016) and adolescent girls and young women (AGYW) (Dellar et al., 2015), continue to face disproportionately high HIV burdens, highlighting the necessity of integrated and targeted national policies. The effectiveness of national AIDS programmes (NAPs), particularly the delivery of essential services like HIV prevention for sex workers, is often contingent upon the formality of national implementation guidelines (Wilson, 2015) and robust coordination with the broader social protection system (UNAIDS, 2012). While the relationship between policy adoption and health outcomes is conceptually clear (e.g., Camilletti, 2020), empirical evidence linking the reported existence of specific national structural policies, instead of the year in which those policies were introduced, to the provision of key HIV services in low- and middle-income settings remains scarce (Gruskin et al., 2013). This study addresses this research gap by focusing on Southeast Asia, a region with varied policy landscapes and HIV epidemic profiles.

HIV epidemics in this region are largely concentrated among key populations, and the legal status of sex work is considerably different across countries. For sex workers in these settings, exclusion from social protection is both administrative and legal. Eligibility for cash transfers, contributory health insurance, and disability benefits is commonly conditional upon national identity documentation, registered residence, or evidence of formal employment. Sex workers are often unable to satisfy these conditions, so HIV-related support cannot reach them through mainstream social protection channels.

We hypothesise that such exclusion affects the presence of targeted HIV prevention services in two ways over time. First, where mainstream social protection excludes sex workers, a separate and targeted HIV prevention component is necessary for HIV-related support to reach this population, so the contemporaneous association between reported barriers and a reported prevention component is expected to be positive. Second, because such a targeted component operates outside the social protection system and lacks any statutory financing, the continued presence of barriers is expected to be associated with the subsequent absence of that component. Utilising a panel dataset of eight Southeast Asian countries spanning 2018–2025, this study aims to quantify the within-country associations between five national policy indicators (note: excluding the outcome indicator: the HIV prevention service component targeted at sex workers)—ranging from social protection coordination to the existence of legal barriers—and the presence of HIV prevention service components specifically targeting sex workers.

## Methods

### Data Collection and Variables

The primary source for the panel data is the UNAIDS Global AIDS Monitoring (GAM) framework, specifically through data collected via the National Commitments and Policy Instrument (NCPI) surveys. We downloaded all available data covering HIV-related laws and policies from the UNAIDS data repository. We performed data collection, restructuring, filtering, and imputation using Python within the Cursor environment.

Countries were eligible for inclusion if they reported in two or more years. Brunei and Vietnam each reported in a single year and were, therefore, excluded. The FE model measures how the outcome changes within each country over time. A country observed in only one year has no change to measure, and, thus, cannot contribute to the FE estimation. The resulting eight Southeast Asian country panel was extracted for statistical data analysis (Appendix 1). We harmonised the six selected policy indicators into dummies for panel regression.

Four indicators were already recorded as binary and were coded as 1 where the government reported the policy or barrier to be present and 0 where the government reported the policy or barrier to be absent. The remaining two indicators, being the social protection coordination mechanism and national SOPs for HIV prevention programmes for sex workers, were recorded in three categories, and we recoded each of them as 1 for the highest policy level and 0 for the partial or low policy level or where no corresponding policy exists. We transformed the non-dummy data into binary variables, allowing the models to measure the impact of each specific policy level (i.e., high policy level) against a designated baseline (i.e., partial/low policy level or no corresponding policy exists). To address the challenge of intermittent reporting over time, we applied forward filling (i.e., Last Observation Carried Forward (LOCF)), which assumes a policy’s status remains constant in subsequent non-reporting years (Lachin, 2016). We structured the final dataset as a balanced panel covering the period from 2018 to 2025.

The dependent variable measures the presence (1) or absence (0) of a key HIV prevention service component targeted at sex workers, as reported by the government. The dependent variable records what the national HIV response is reported to contain, and does not measure the coverage, quality, or accessibility of the HIV prevention service component targeted at sex workers.

One of the explanatory variables measures the existence (1) or non-existence (0) of legal or administrative barriers that prevent people living with HIV and key populations, including sex workers, from accessing social protection. The variable assesses the presence of negative structural impediments, but not the introduction of those impediments, and describes the legal and administrative environment that the government has adopted. The remaining four variables measure the adoption of programmatic policies. These are the presence (1) or absence (0) of the highest level of social protection coordination mechanism, meaning a mechanism that includes NAP representatives; the presence (1) or absence (0) of the highest level of formality for implementation, meaning whether national SOPs apply for all organisations providing HIV prevention services to sex workers; the adoption (1) or non-adoption (0) of a national policy that acknowledges and aims to mitigate the burden of unpaid care work related to HIV; and the adoption (1) or non-adoption (0) of a policy that explicitly targets AGYW for social protection benefits. Appendix 2 shows the concise description of the list of variables used in this study.

H2>Statistical Analysis

As the dependent variable is binary, we estimated linear probability models with country Fixed Effects (FE). The FE estimator removes each country’s time-invariant characteristics by subtracting each country’s mean values from that country’s own observations, so the reported associations are identified from change within countries over the study period but not from differences between countries. We estimated RE models for comparison. We note that the FE and Random-Effects (RE) terminology used here follows panel econometrics, in that the FE estimator is a within-country transformation and not a common-effect assumption. We selected between the two estimators using the Sargan-Hansen C-statistic, being the robust form of the Hausman test appropriate under heteroscedasticity, which tests whether the country-specific effects are correlated with the explanatory variables. Standard errors are clustered at the country level throughout, so as to allow for autocorrelation and heteroscedasticity within countries. We assessed linear dependence between the explanatory variables using Spearman rank correlations and Variance Inflation Factors (VIF). Model equations and estimation assumptions are set out in full in Appendix 4.

Reliance on a single estimation model (Table 3) would not adequately demonstrate the robustness of these estimates. We built an additional robustness check (Table 4) by re-running the regression using lagged explanatory variables, testing whether policy effects exist with a one-period time lag. We selected a one-period lag because the NCPI records policy conditions annually, so a policy condition reported in one year is the most proximate plausible influence on the HIV prevention service component reported in the following year.

### Ethics Statement

Ethics approval was not required for this study as all data were drawn from publicly available, anonymised, aggregated surveillance datasets published by the UNAIDS GAM framework.

### Patient and Public Involvement Statement

As indicated, the data used for this study is a secondary, aggregated panel dataset, comprising national policy and commitment indicators reported by governments and national HIV programmes. The development of our research question—analysing the impact of structural policies on HIV prevention—was informed by these national-level policy priorities and outcomes, which indirectly suggests patient needs for comprehensive and barrier-free services. Consequently, the research design and conduct did not involve direct interaction, recruitment, or intervention with individual patients or study participants, as the study relies entirely on publicly available data. The results of this work will be disseminated through academic publication and any subsequent presentation, which serves as a primary link to patient and community advocacy groups.

## Results

The mean values in Table 1 describe the distribution of national HIV policy adoption across the eight Southeast Asian countries within the panel data (2018–2025). The policies demonstrating the highest adoption rates (mean ≥ 0.60) are those concerning specific populations and coordination mechanisms. Specifically, the most common policies practised in Southeast Asia are the ones recognising AGYW as key beneficiaries of social protection (*sp_agywi,t* ≈ 0.63) and upholding social protection coordination mechanism/platform which includes the NAP (*sp_coord_napi,t* ≈ 0.63). The least adopted policy within the region is the presence of national SOPs for HIV prevention programmes for sex workers (*national_sopi,t* ≈ 0.16).

**Table 1:** Summary of All Variables.

| Variable | Observations | Mean | S.D. | Min. | Max. |
| --- | --- | --- | --- | --- | --- |
| $country_i$ | 64 | 4.5 | 2.31 | 1 | 8 |
| $year_t$ | 64 | 2021.5 | 2.31 | 2018 | 2025 |
| $hiv\_prevention\_sex\_workers_{i,t}$ | 64 | 0.45 | 0.50 | 0 | 1 |
| $barriers\_sp_{i,t}$ | 64 | 0.45 | 0.50 | 0 | 1 |
| $sp\_upaid\_care\_hiv_{i,t}$ | 64 | 0.39 | 0.49 | 0 | 1 |
| $sp\_agyw_{i,t}$ | 64 | 0.63 | 0.49 | 0 | 1 |
| $sp\_coord\_nap_{i,t}$ | 64 | 0.63 | 0.49 | 0 | 1 |
| $national\_sop_{i,t}$ | 64 | 0.16 | 0.37 | 0 | 1 |

Multicollinearity between independent variables was not problematic in this study, as shown by both the Spearman Rank Correlation Table (Table 2) and the VIF results (Appendix 3). In Table 2, the highest correlations observed among the independent variables are 0.6000 (between *sp_agywi,t* and *sp_coord_napi,t*; *p* < 0.001) and 0.4217 (between *sp_upaid_care_hivi,t* and *sp_coord_napi,t*; *p* < 0.001). Since no correlation coefficient approaches the critical threshold of ≥ 0.80, no extreme linear relationships exist between the predictors. Furthermore, the analysis of multicollinearity in Appendix 3 shows a Mean VIF of 1.37, and all individual VIF values are well below the threshold of five—meaning the VIF values are conventionally accepted. These low VIF scores affirm that the explanatory variables are not excessively correlated with one another, so as to ensure that the estimated regression coefficients are reliable and interpretable.

**Table 2:** Spearman Rank Correlation Table between Independent Variables.

|  | *barriers_sp<sub>i,t</sub>* | *sp_unpaid_care_hiv<sub>i,t</sub>* | *sp_agyw<sub>i,t</sub>* | *sp_coord_nap<sub>i,t</sub>* | *national_sop<sub>i,t</sub>* |
| --- | --- | --- | --- | --- | --- |
| *barriers_sp<sub>i,t</sub>* | 1.0000 |  |  |  |  |
| *sp_unpaid_care_hiv<sub>i,t</sub>* | -0.0854 | 1.0000 |  |  |  |
| *sp_agyw<sub>i,t</sub>* | -0.0081 | 0.3556** | 1.0000 |  |  |
| *sp_coord_nap<sub>i,t</sub>* | 0.0567 | 0.4217*** | 0.6000*** | 1.0000 |  |
| *national_sop<sub>i,t</sub>* | 0.2999* | 0.0965 | 0.0667 | 0.1556 | 1.0000 |
| N (Observations) | 64 |  |  |  |  |

Note: \*\*\*, \*\*, \* denotes significance at the 0.1%, 1%, and 5% level, respectively.

**Table 3:**
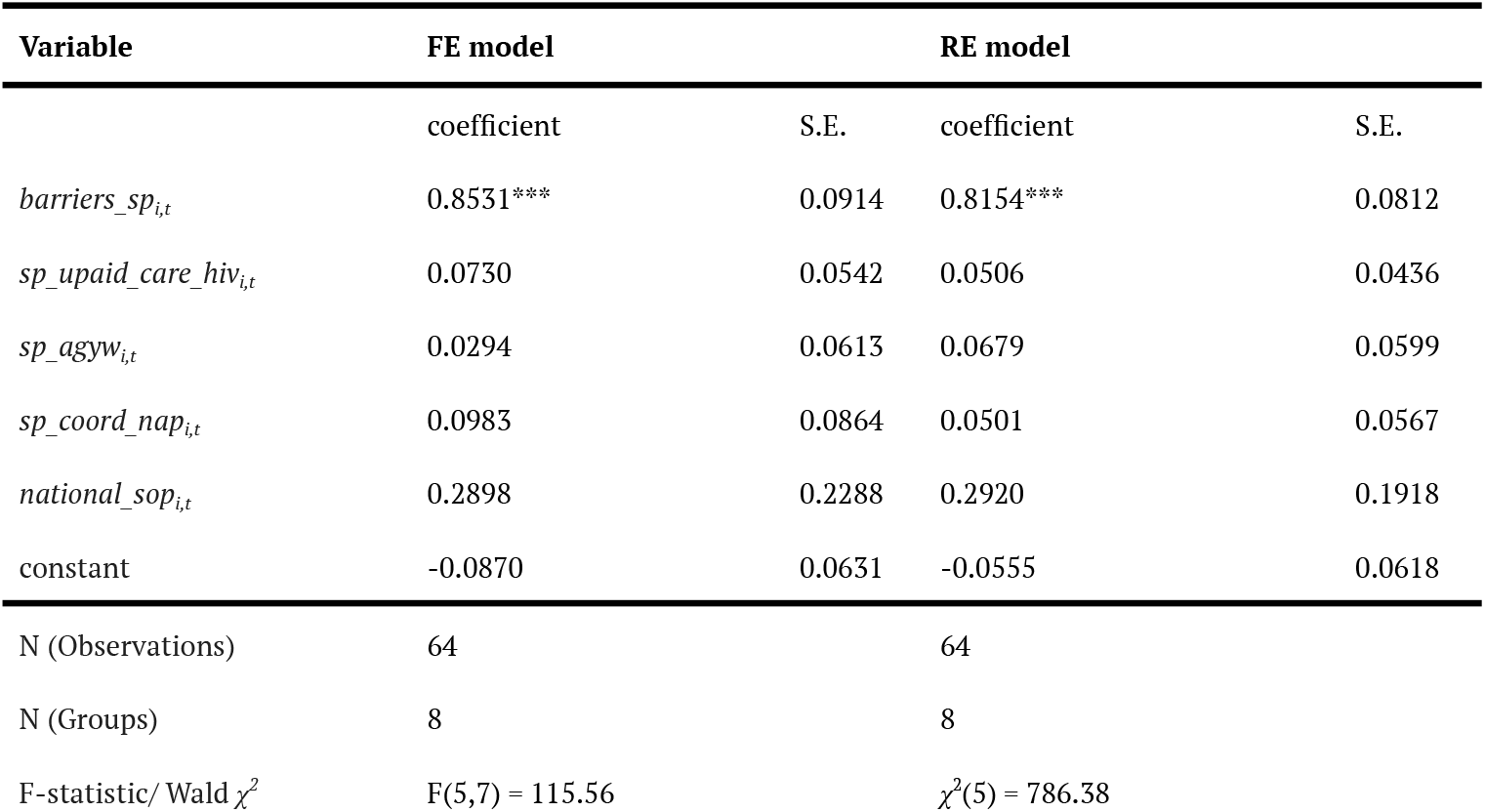
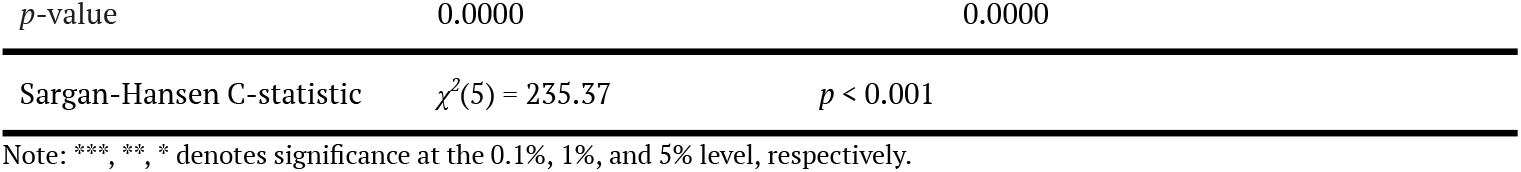
Regression Results for HIV Prevention Service for Sex Workers (FE vs. RE with Robust Specification Test)

**Table 4:** Regression Results for HIV Prevention Service for Sex Workers (Lagged ExplanatoryVariables)

| Variable | FE model |  | RE model |  |
| --- | --- | --- | --- | --- |
|  | coefficient | S.E. | coefficient | S.E. |
| *barriers_sp<sub>i,t-1</sub>* | -0.3540* | 0.1409 | -0.1280 | 0.1293 |
| *sp_unpaid_care_hiv<sub>i,t-1</sub>* | -0.0434 | 0.3706 | -0.1229 | 0.1559 |
| *sp_agyw<sub>i,t-1</sub>* | -0.0371 | 0.2584 | -0.0263 | 0.1970 |
| *sp_coord_nap<sub>i,t-1</sub>* | 0.0013 | 0.3018 | 0.1654 | 0.1789 |
| *national_sop<sub>i,t-1</sub>* | -0.2053 | 0.1376 | -0.3073* | 0.1221 |
| constant | 0.7344* | 0.2184 | 0.5572*** | 0.1361 |
| N (Observations) | 56 |  | 56 |  |
| N (Groups) | 8 |  | 8 |  |
| F-statistic/ Wald $\chi^2$ | F(5,7) = 6.60 | | $\chi^2(5) = 50.60$ | |
| p-value | 0.0140 |  | 0.0000 |  |
| $\sigma_u = 0$ | | | | |

Note: \*\*\*, \*\*, \* denotes significance at the 0.1%, 1%, and 5% level, respectively.

Table 3 presents the initial regression results using contemporaneous policy variables. We selected between the FE and RE models as described in the Methods. The Sargan-Hansen C-statistic is *χ*^2^(5) = 235.37 (*p* < 0.001). We, therefore, reject the null hypothesis that the country-specific effects are uncorrelated with the explanatory variables. This means the RE estimator is inconsistent, and we, thus, report the FE estimator as the primary specification. Table 3 shows that we are statistically confident (*p* < 0.001) to argue that the reported existence of barriers to accessing social protection is positively associated with the reported presence of an HIV prevention service component targeted at sex workers, within countries and over time (*β* = 0.8531). In descriptive terms, the HIV prevention service component targeted at sex workers was reported in 93.1% of country-years in which barriers to social protection were reported, against 5.7% of country-years in which no such barriers to social protection were reported.

Table 4 provides the results using one-period lagged explanatory variables as an important robustness check. For the model selection, the RE model output in Table 4 includes *σ*_*u*_ = 0, signalling a failure to estimate the variance of the unobserved country effect (*α*_*i*_), which reinforces the reliability of the FE model. In the FE model with lagged variables, we are statistically confident (*p* = 0.011) to argue that the previous year’s reported barriers to social protection are negatively associated with the HIV prevention service component targeted at sex workers reported in the current year (*β* = −0.3540). In descriptive terms, the HIV prevention service component targeted at sex workers was reported in 39.3% of country-years following a reported barrier to social protection, against 57.1% of country-years following no reported barrier to social protection.

## Discussion

The findings of this study provide new quantitative evidence on the association between national structural policy and the reported HIV prevention service component for sex workers in Southeast Asia. Our primary finding from the FE model is the statistically significant and positive contemporaneous association between the existence of legal or administrative barriers to social protection in a country and the presence of HIV prevention services for sex workers in the same country. While seemingly counterintuitive, this result might be explained by the necessity of targeted provision where mainstream social protection excludes sex workers.

Where legal or administrative barriers prevent sex workers from accessing social protection, HIV-related support cannot reach them through mainstream channels, and a separate prevention component targeted at sex workers becomes necessary. Countries that do not report legal or administrative barriers to social protection may serve sex workers through general social protection provision, and may, therefore, have no HIV prevention service component targeted at sex workers to report. This interpretation aligns with the first hypothesis presented in the Introduction, and suggests the contemporaneous association shows the substitution of targeted provision for mainstream provision, but not the effectiveness of national HIV programmes.

On the contrary, our robustness check using lagged variables revealed that the previous year’s reported barriers were negatively and statistically significantly associated with the HIV prevention service component targeted at sex workers reported in the current year. This aligns with the second hypothesis presented in the Introduction. A targeted component that exists because the social protection system excludes sex workers operates outside that system and lacks any statutory financing, and the continued presence of barriers to social protection is associated with the HIV prevention service component targeted at sex workers no longer being reported in the following year. We note that the lagged association reported in Table 4 holds between two reported policy indicators, and does not establish that structural barriers reduce the effectiveness or sustainability of targeted HIV-related health programmes for sex workers.

In practice, the barriers recorded by *barriers_sp* are both administrative and legislative. Sex work is criminalised in all eight countries in this panel, which itself removes sex workers from the labour protections and contributory social security schemes available to formally employed workers (Godwin, 2012). Administrative conditions compound this exclusion. In Malaysia, access to services is restricted by difficulties in obtaining identity cards, and in Laos internal migrant sex workers face difficulties in accessing services because of residency registration requirements (Godwin, 2012). Thailand is a partial exception, in that sex workers may access some entitlements under the state social security scheme where contributions are made, although the *Labour Protection Act* is not enforced in this sector (Godwin, 2012).

The opposite directions of the contemporaneous and lagged associations have not previously been documented using panel data methods in this regional context. Gruskin et al. (2013), in an analysis of National Composite Policy Index data across multiple countries, showed that the national policy environment is heterogeneously linked to HIV service outcomes, which aligns with our finding that policy indicators do not uniformly predict the reported HIV prevention service component across country-years. Wilson (2015) demonstrated that the effectiveness of HIV prevention programmes for sex workers depends substantially on the formalisation of national implementation guidelines, which is a mechanism featured in our study through the national SOP variable, although this coefficient did not reach statistical significance in our sample. The negative lagged effect further supports Nkengasong and Ratevosian’s (2023) argument that legal and policy barriers constitute a structural impediment to the HIV/AIDS response over time. In sum, these empirical results distinguish short-term policy recognition from long-term policy reform, which are associated in opposite directions with the reported HIV prevention service component for sex workers.

A key limitation of this study is that both the dependent variable and the explanatory variables are measures of what governments report about their own policies, not measures of the health services that populations actually use. The dependent variable measures whether a government reported that an HIV prevention service component targeted at sex workers forms part of its national HIV response. The dependent variable does not measure service accessibility, coverage, or uptake. An association between the dependent variable and reported barriers to social protection, therefore, describes what governments report, and does not describe what sex workers receive. Second, FE estimation removes country characteristics that do not change over the study period, but FE estimation does not control for confounders that do change over the study period. Changes in external financing, including Global Fund and PEPFAR transitions, disruption to health services during the COVID-19 pandemic, and revisions to the NCPI reporting instrument all changed over 2018 to 2025 and are not accounted for in our models. Third, the lag structure cannot establish which of the two reported indicators affects the other. Lagging the explanatory variables by one year means that reported barriers to social protection are measured in the year before the HIV prevention service component targeted at sex workers.

Measuring the barriers a year earlier does not show that reported barriers affect the HIV prevention service component targeted at sex workers, because we would find the same association if the HIV prevention service component targeted at sex workers reported in one year affected reported barriers to social protection in the following year. Fourth, the models are linear probability models, in which ordinary least squares is applied to a binary dependent variable. The predicted probabilities from a linear probability model may fall below zero or exceed one, which is not possible for a probability, and this is a recognised limitation of linear probability models. Finally, this study relies on policy self-reporting through the UNAIDS GAM framework, which measures policy adoption and not the quality or scale of policy implementation, and the dependent variable does not record who funds the HIV prevention service component targeted at sex workers. Future research should incorporate sub-national data or policy implementation scores.

## Conclusions

Using panel data covering eight Southeast Asian countries over 2018 to 2025, this study finds that the relationship between structural barriers to social protection and the reported HIV prevention service component for sex workers is not uniform across time. The contemporaneous association is positive and statistically significant, indicating that countries that have reported the existence of legal or administrative barriers to social protection are, in the same year, more likely to report an HIV prevention service component targeting sex workers. The lagged association runs in the opposite direction, in which the prior year’s presence of those same barriers is negatively and significantly associated with the HIV prevention service component targeting sex workers reported in the current year. These results indicate that the formal recognition of social protection barriers may occur alongside early investment in HIV prevention services, but that barriers left unaddressed are associated with the absence of the HIV prevention service component targeting sex workers in the following year.

Our findings contribute empirically to the panel data literature from an underrepresented region on structural HIV policy. This study reinforces the argument, aligning with the UNAIDS Global AIDS Strategy 2021 to 2026, that the removal of structural barriers is important for sustained HIV prevention service delivery. National HIV programmes in Southeast Asia should treat barrier removal as a time-sensitive operational priority, integrated into annual programme reviews but not addressed as a long-term strategic aspiration. Future research should extend this approach to other low- and middle-income regions and incorporate implementation-quality measures to distinguish between policy adoption and effective policy execution.

## Appendix 1: List of Relevant Variables

| *country<sub>i</sub>* | *year<sub>t</sub>* | *hiv_prevention_sex_workers<sub>i,t</sub>* | *barriers_sp<sub>i,t</sub>* | *sp_upaid_care_hiv<sub>i,t</sub>* | *sp_agyw<sub>i,t</sub>* | *sp_coord_nap<sub>i,t</sub>* | *national_sop<sub>i,t</sub>* |
| --- | --- | --- | --- | --- | --- | --- | --- |
| Cambodia | 2018 | 1 | 1 | 0 | 1 | 0 | 0 |
| Cambodia | 2019 | 1 | 1 | 0 | 1 | 1 | 0 |
| Cambodia | 2020 | 0 | 0 | 0 | 1 | 1 | 0 |
| Cambodia | 2021 | 1 | 1 | 0 | 1 | 1 | 1 |
| Cambodia | 2022 | 1 | 1 | 0 | 1 | 1 | 1 |
| Cambodia | 2023 | 0 | 0 | 0 | 0 | 1 | 0 |
| Cambodia | 2024 | 1 | 1 | 0 | 0 | 1 | 1 |
| Cambodia | 2025 | 0 | 0 | 0 | 1 | 1 | 0 |
| Indonesia | 2018 | 0 | 0 | 0 | 1 | 1 | 0 |
| Indonesia | 2019 | 0 | 0 | 0 | 1 | 1 | 0 |
| Indonesia | 2020 | 0 | 0 | 0 | 0 | 0 | 0 |
| Indonesia | 2021 | 1 | 1 | 0 | 0 | 1 | 0 |
| Indonesia | 2022 | 1 | 1 | 0 | 0 | 0 | 0 |
| Indonesia | 2023 | 0 | 0 | 0 | 0 | 0 | 0 |
| Indonesia | 2024 | 1 | 1 | 0 | 0 | 0 | 0 |
| Indonesia | 2025 | 0 | 0 | 0 | 0 | 0 | 0 |
| Laos | 2018 | 1 | 1 | 1 | 1 | 1 | 0 |
| Laos | 2019 | 1 | 1 | 1 | 1 | 1 | 0 |
| Laos | 2020 | 1 | 1 | 1 | 1 | 1 | 0 |
| Laos | 2021 | 1 | 1 | 0 | 1 | 1 | 0 |
| Laos | 2022 | 1 | 1 | 0 | 1 | 1 | 0 |
| Laos | 2023 | 0 | 0 | 0 | 1 | 0 | 0 |
| Laos | 2024 | 1 | 1 | 0 | 0 | 0 | 1 |
| Laos | 2025 | 0 | 0 | 1 | 1 | 0 | 0 |
| Malaysia | 2018 | 0 | 0 | 0 | 0 | 1 | 0 |
| Malaysia | 2019 | 1 | 1 | 0 | 1 | 1 | 0 |
| Malaysia | 2020 | 0 | 0 | 0 | 1 | 1 | 0 |
| Malaysia | 2021 | 1 | 1 | 0 | 1 | 1 | 0 |
| Malaysia | 2022 | 0 | 1 | 0 | 0 | 0 | 0 |
| Malaysia | 2023 | 0 | 1 | 0 | 0 | 0 | 0 |
| Malaysia | 2024 | 1 | 1 | 0 | 0 | 0 | 0 |
| Malaysia | 2025 | 1 | 1 | 0 | 0 | 0 | 0 |
| Myanmar | 2018 | 0 | 0 | 0 | 1 | 0 | 0 |
| Myanmar | 2019 | 1 | 1 | 0 | 1 | 0 | 1 |
| Myanmar | 2020 | 0 | 0 | 0 | 0 | 0 | 0 |
| Myanmar | 2021 | 0 | 0 | 0 | 0 | 0 | 0 |
| Myanmar | 2022 | 0 | 0 | 0 | 0 | 0 | 0 |
| Myanmar | 2023 | 0 | 0 | 0 | 0 | 0 | 0 |
| Myanmar | 2024 | 1 | 1 | 0 | 0 | 0 | 0 |
| Myanmar | 2025 | 0 | 0 | 0 | 0 | 0 | 0 |
| Philippines | 2018 | 0 | 0 | 0 | 1 | 1 | 0 |
| Philippines | 2019 | 1 | 1 | 1 | 1 | 1 | 0 |
| Philippines | 2020 | 0 | 0 | 1 | 1 | 1 | 0 |
| Philippines | 2021 | 1 | 1 | 1 | 1 | 1 | 0 |
| Philippines | 2022 | 1 | 1 | 1 | 1 | 1 | 0 |
| Philippines | 2023 | 0 | 0 | 0 | 1 | 1 | 0 |
| Philippines | 2024 | 1 | 1 | 1 | 1 | 1 | 0 |
| Philippines | 2025 | 0 | 0 | 1 | 1 | 1 | 0 |
| Singapore | 2018 | 0 | 0 | 0 | 1 | 0 | 0 |
| Singapore | 2019 | 1 | 0 | 1 | 1 | 1 | 1 |
| Singapore | 2020 | 0 | 0 | 1 | 1 | 1 | 0 |
| Singapore | 2021 | 0 | 0 | 1 | 1 | 1 | 0 |
| Singapore | 2022 | 0 | 0 | 1 | 1 | 1 | 0 |
| Singapore | 2023 | 0 | 0 | 1 | 1 | 1 | 0 |
| Singapore | 2024 | 1 | 0 | 1 | 1 | 1 | 1 |
| Singapore | 2025 | 0 | 0 | 1 | 1 | 1 | 0 |
| Thailand | 2018 | 0 | 0 | 1 | 0 | 0 | 0 |
| Thailand | 2019 | 0 | 0 | 1 | 0 | 0 | 0 |

## Appendix 2: Description of Variable Definitions

| Variable | Variable Label | Description (Policy Interpretation) |
| --- | --- | --- |
| *hiv_prevention_sex_workers<sub>i,t</sub>* | Including any prevention strategy for sex workers, selected countries | This variable measures the presence (1) or absence (0) of a key HIV prevention service component targeted at sex workers. This is the dependent variable of this research. |
| *barriers_sp<sub>i,t</sub>* | Barriers to access social protection, selected countries | This variable measures the existence (1) or non-existence (0) of legal or administrative barriers that prevent vulnerable people (like people living with HIV) from accessing social protection. The variable assesses the presence of negative structural impediments. |
| *sp_unpaid_care_hiv<sub>i,t</sub>* | Social protection strategy/ policy/ framework: Addresses unpaid care work in the HIV context, selected countries | This measures the adoption (1) or non-adoption (0) of a national policy that specifically acknowledges and aims to mitigate the burden of unpaid care work related to HIV. |
| *sp_agyw<sub>i,t</sub>* | Social protection strategy/ policy/ framework: Recognises AGYW as key beneficiaries, selected countries | This variable measures the adoption (1) or non-adoption (0) of a policy that explicitly targets AGYW for social protection benefits. The variable assesses the policy reach of targeted inclusion. |
| *sp_coord_nap<sub>i,t</sub>* | Social protection coordination mechanism/platform, selected countries | This is a dummy policy variable (1/0) created from a raw multi-categorical indicator. This variable measures the presence (1) or absence (0) of the highest level of social protection coordination mechanism, meaning a mechanism that includes NAP representatives to ensure the integration and effective coordination of social protection efforts with the national HIV response. |
| *national_sop<sub>i,t</sub>* | SOPs or national implementation guidelines for HIV prevention programmes for sex workers, selected countries | This is a dummy policy variable (1/0) created from a raw multi-categorical indicator. This variable measures the presence (1) or absence (0) of the highest level of formality for the implementation of HIV prevention programmes for sex workers, meaning whether national SOPs apply for all organisations providing HIV prevention services to sex workers. |

## Appendix 3: Variance Inflation Factor (VIF)

| Variable | VIF | 1/VIF |
| --- | --- | --- |
| *barriers_sp<sub>i,t</sub>* | 1.12 | 0.89 |
| *sp_upaid_care_hiv<sub>i,t</sub>* | 1.27 | 0.79 |
| *sp_agyw<sub>i,t</sub>* | 1.60 | 0.63 |
| *sp_coord_nap<sub>i,t</sub>* | 1.73 | 0.58 |
| *national_sop<sub>i,t</sub>* | 1.13 | 0.89 |
| Mean VIF | 1.37 |  |

## Appendix 4

### Modelling

When building linear regression models for panel data, we used *hiv_prevention_sex_workersi,t* as the dependent variable and *barriers_spi,t, sp_upaid_care_hivi,t, sp_agywi,t, sp_coord_napi,t* and *national_sopi,t* as the independent variables (in Table 3). Here are the regression equations and assumptions for the FE and Random-Effects (RE) models:

#### (1) FE Model

The FE model (or “within” estimator) controls for all unobserved differences between countries by removing the country-specific mean, so as to isolate the effect of changes within each country over time.

(1)FE Model: *hiv_prevention_sex_workers*_*i,t*_ *= β*_*0*_ *+ β*_*1*_*barriers_sp*_*i,t*_ *+ β*_*2*_*sp_upaid_care_hiv*_*i,t*_ *+ β*_*3*_*sp_agyw*_*i,t*_ *+ β*_*4*_*sp_coord_nap*_*i,t*_ *+ β*_*5*_*national_sop*_*i,t*_ *+ α*_*i*_ *+ ϵ*_*it*_

Assumptions for the FE estimation:

1. **Strict exogeneity**: The time-varying error term c_it_ is uncorrelated with the regressors in all time periods: *E[ϵ*_*it*_ | *X*_*i1*_, …, *X*_*iT*_, *α*_*i*_] = 0.
2. **No perfect collinearity:** The time-varying regressors (X_it_) must vary over time within each country.
3. **Error structure**: The idiosyncratic errors _(cit)_ are assumed to be independent across observations, but because *vce(robust)* was used, the model accommodates heteroscedasticity (non-constant variance) in the errors.

#### (2) RE Model

The RE model assumes that the unobserved country-specific effects (αi) are random and uncorrelated with the predictors. It allows for the estimation of time-invariant variables (Z_i_).

(2)RE Model: *hiv_prevention_sex_workers*_*i,t*_ *= β*_*0*_ *+ β*_*1*_*barriers_sp*_*i,t*_ *+ β*_*2*_*sp_upaid_care_hiv*_*i,t*_ *+ β*_*3*_*sp_agyw*_*i,t*_ *+ β*_*4*_*sp_coord_nap*_*i,t*_ *+ β*_*5*_*national_sop*_*i,t*_ *+ (α*_*i*_ *+ ϵ*_*it*_)

Assumptions for the RE estimation:

1. **No correlation**: The country-specific error term (α_i_) must be uncorrelated with all explanatory variables (X_it_ and Z_i_). This assumption is tested by the Sargan-Hansen C-statistic.
2. **Error structure**: The composite error term (u_it_ = α_i_ + ϵ_it_) is assumed to be independent across countries.
3. **Heteroscedasticity**: As *vce(robust)* was used, the estimation accommodates heteroscedasticity in the errors (ϵ_it_ and α_i_).

#### (3) FE Model (Lagged)

The FE model removes all time-invariant country heterogeneity (α_i_). It tests how changes in the lagged explanatory variables within a country affect the current outcome.

(3)FE Model: *hiv_prevention_sex_workers*_*i,t*_ *= β*_*0*_ *+ β*_*1*_*barriers_sp*_*i,t-1*_ *+ β*_*2*_*sp_upaid_care_hiv*_*i,t-1*_ *+ β*_*3*_*sp_agyw*_*i,t-1*_ *+ β*_*4*_*sp_coord_nap*_*i,t-1*_ *+ β*_*5*_*national_sop*_*i,t-1*_ *+ α*_*i*_ *+ ϵ*_*it*_

Assumptions for the FE estimation:

1. **Strict exogeneity (lagged)**: The idiosyncratic error term (ϵ_it_) is uncorrelated with the lagged regressors (X_1,t-1_) across all time periods.
2. **No perfect collinearity**: The lagged regressors (X_1,t-1_) must vary over time within each country.
3. **Time-invariant variable omission**: The time-invariant variable (Z_i_) is absorbed into α_i_ and is omitted from the estimation.
4. **Error structure (clustered)**: The use of *vce(cluster country_id)* ensures that the inference is valid, as it explicitly assumes:
  a. Errors are correlated within each country (autocorrelation is allowed).
  b. Errors may have different variances across countries (heteroscedasticity is allowed).
  c. Errors are independent between countries (clusters).

#### RE Model (Lagged)

The RE model estimates the effect using a weighted average of the within- and between-country variation. It allows for the estimation of the time-invariant variable (Z_i_).

(4)RE Model: *hiv_prevention_sex_workers*_*i,t*_ *= β*_*0*_ *+ β*_*1*_*barriers_sp*_*i,t-1*_ *+ β*_*2*_*sp_upaid_care_hiv*_*i,t-1*_ *+ β*_*3*_*sp_agyw*_*i,t-1*_ *+ β*_*4*_*sp_coord_nap*_*i,t-1*_ *+ β*_*5*_*national_sop*_*i,t-1*_ *+ (α*_*i*_ *+ ϵ*_*it*_)

Assumptions for the RE estimation:

1. **No correlation:** The country-specific error term (α_i_) must be uncorrelated with all lagged and time-invariant explanatory variables (X_1,t-1_ and Z_i_).
2. **Lag structure**: It assumes the relationship between the predictors and the outcome is fully captured by the one-period lag structure.
3. **Error structure (clustered)**: Similar to the FE model, *vce(cluster country_id)* provides robust inference by allowing for autocorrelation and heteroscedasticity within each country cluster.

## Acknowledgements

N/A.

## Author Contributions

Conceptualisation: J.H.; Methodology: J.H.; Data curation and formal analysis: J.H.; Writing – original draft: J.H.; Writing – review and editing: J.H. and A.S.; Supervision: A.S.

## Data Availability

The source data used in this study are publicly available from the UNAIDS GAM framework via the UNAIDS data repository at https://aidsinfo.unaids.org/. The constructed panel dataset used for all analyses, covering eight Southeast Asian countries from 2018 to 2025, is reproduced in full in Appendix 1 of this manuscript. No additional data beyond what is presented in Appendix 1 were used in the regression analyses.

